# Immune responses following 3^rd^ and 4^th^ doses of heterologous and homologous COVID-19 vaccines in kidney transplant recipients

**DOI:** 10.1101/2022.04.29.22274396

**Authors:** Tina Thomson, Maria Prendecki, Sarah Gleeson, Paul Martin, Katrina J Spensley, Rute Cardoso De Aguiar, Bynvant Sandhu, Charlotte Seneschall, Jaslyn Gan, Candice L. Clarke, Shanice Lewis, Graham Pickard, David Thomas, Stephen P. McAdoo, Liz Lightstone, Alison Cox, Peter Kelleher, Michelle Willicombe

**Author notes:** Corresponding author: Dr Michelle Willicombe, Centre for Inflammatory Disease Department of Immunology and Inflammation, Faculty of Medicine, Imperial College London, Hammersmith Hospital Campus, London, W12 0NN. These authors contributed equally to this work (Tina Thomson, Maria Prendecki, Michelle Willicombe).

## Abstract

**Background:** Solid organ transplant recipients have attenuated immune responses to SARS-CoV-2 vaccines. In this study, we report on immune responses to 3^rd^- (V3) and 4^th^- (V4) doses of heterologous and homologous vaccines in a kidney transplant population.

**Methods:** 724 kidney transplant recipients were prospectively screened for serological responses following 3 primary doses of a SARS-CoV2 vaccine. 322 patients were sampled post-V4 for anti-spike (anti-S), with 69 undergoing assessment of SARS-CoV-2 T-cell responses. All vaccine doses were received post-transplant, only mRNA vaccines were used for V3 and V4 dosing. All participants had serological testing performed post-V2 and at least once prior to their 1^st^ dose of vaccine.

**Results:** 586/724 (80.9%) patients were infection-naïve post-V3; 141/2586 (24.1%) remained seronegative at 31 (21-51) days post-V3. Timing of vaccination in relation to transplantation, OR: 0.28 (0.15-0.54), p=0.0001; immunosuppression burden, OR: 0.22 (0.13-0.37), p<0.0001, and a diagnosis of diabetes, OR: 0.49 (0.32-0.75), p=0.001, remained independent risk factors for non-seroconversion. Seropositive patients post-V3 had greater anti-S if primed with BNT162b2 compared with ChAdOx1, p=0.001.

Post-V4, 45/239 (18.8%) infection-naïve patients remained seronegative. De novo seroconversion post-V4 occurred in 15/60 (25.0%) patients who were seronegative post-V3. There was no difference in anti-S post-V4 by vaccine combination, p=0.50. Anti-S post-V4 were sequentially greater in those seroconverting post V2- compared with V3-, and V3- compared with V4-, at 1561 (567-5211), 379 (101-851) and 19 (9.7-48) BAU/ml respectively.

T-cell responses were poor, with only 11/54 (20.4%) infection-naive patients having detectable T-cell responses post-V4, with no difference seen by vaccine type.

**Conclusion:** A significant proportion of transplant recipients remain seronegative following 3- and 4- doses of SARS-CoV-2 vaccines, with poor T-cell responses, and are likely to have inadequate protection against infection.

---

There will now be significant heterogeneity in the immune repertoire against COVID-19 in the population, reflecting a combination of infection due to an array of different variants and evolving vaccination policies over the last 2 years^1^. In the general population, additional booster vaccinations have served to ensure adequate protection against severe infection with the emergence of the Omicron variant. The immune signature against COVID-19 in immunocompromised people could be considered even more diverse than that of the general population, with vaccine responses being additionally dependent upon the underlying condition and treatment^2,3^. Recognised as having attenuated immune responses to COVID-19 vaccines, immunocompromised people in the UK are now being offered their 5^th^ vaccine dose, coupled with eligibility for community therapeutic interventions should they become infected^2,4^.

Some immunocompromised individuals will fail to mount an immune response to vaccination, but there remains no policy for the clinical testing of vaccine responses in this or wider population, which is largely due to the lack of a definition of an adequate response, or correlate of protection. Herein we report on the immune responses to 3^rd^- and 4^th^-doses of heterologous and homologous vaccines in a kidney transplant population, to inform the immune landscape in this severely immunosuppressed population prior to their 5^th^ dose.

## Results

We assessed 724 kidney transplant recipients following 3^rd^ vaccine doses (V3); 586 (80.9%) were infection-naïve, with 138 (19.1%) having evidence of prior infection, *Supplemental Information*, **Figure S1**. A further 322 patients were sampled following a 4^th^ vaccine dose; 239 (74.2%) were infection-naïve, and 83 (25.8%) infection-exposed.

### Serological responses in infection-naïve patients post-V3 and V4

Following V3, 141 (24.1%) infection-naïve patients remained seronegative after a median time of 31 (21-51) days. De novo seroconversion post-V3 occurred in 138/279 (49.5%) patients who were seronegative post-V2, **Figure 1**. Patients who were seropositive post-V3 were more likely to have received the 1^st^ dose of vaccine more than one-year post-transplant (p<0.001), be maintained on tacrolimus monotherapy (p<0.001), primed (V1 and V2) with BNT162b2^2^ (p=0.0086) and not have a diagnosis of diabetes (p=0.005), *Supplemental Information*, **Table S1**. Sampling of seropositive infection-naïve patients post-V3 occurred significantly later than seronegative patients, at a median time of median 33 (21-53) and 24 (21-43) days respectively, p=0.007.

**Figure 1.**
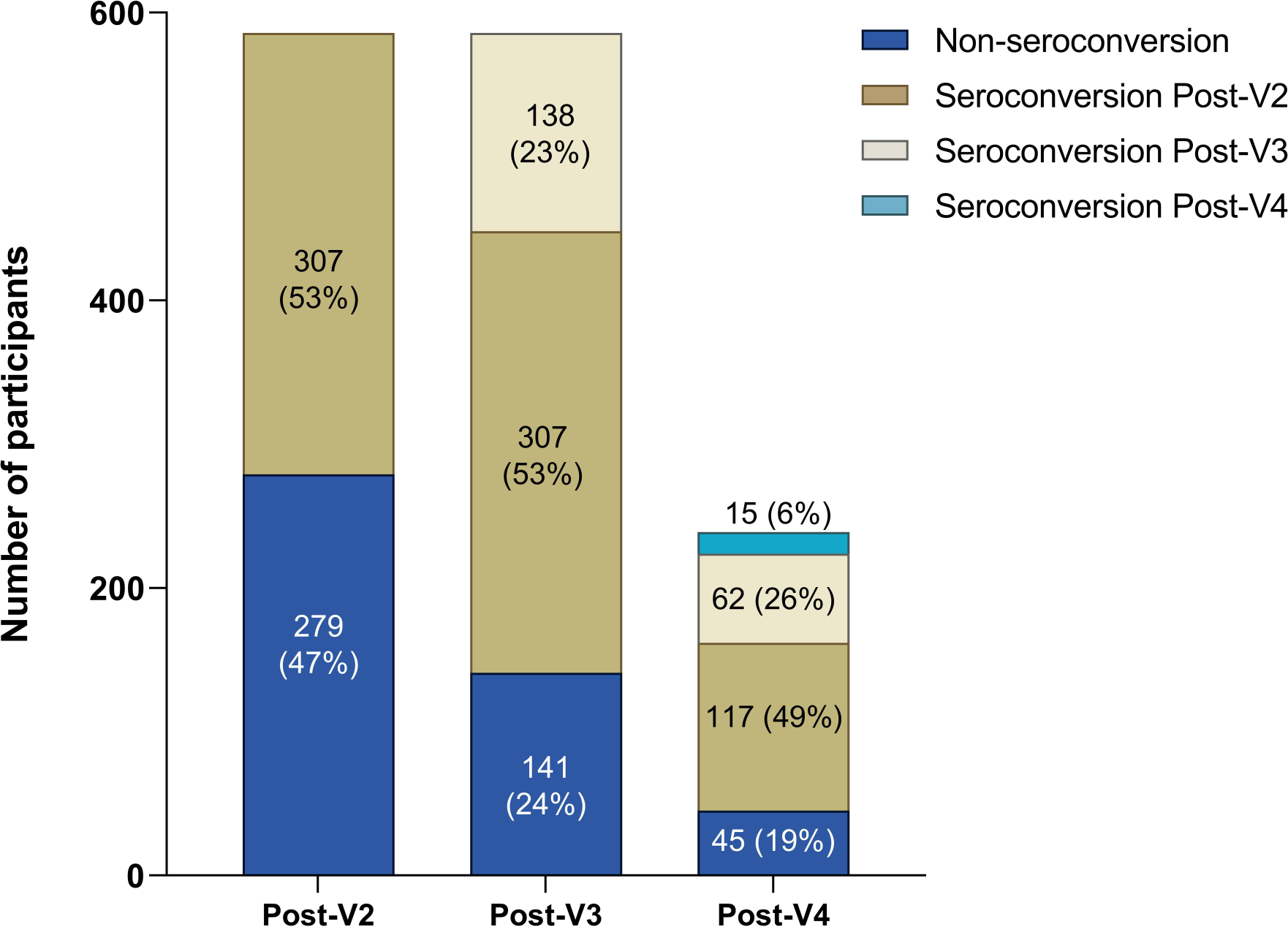
Distribution of serostatus by vaccination dose in infection-naïve individuals. Representative bar chart showing 297 (47%) of patients failed to seroconvert post-V2, 138 (23%) of all patients or 49.5% seronegative patients post-V2, newly seroconverted post-V3. 15/60 (25%) of seronegative patients post-V3, converted post V4.

On multivariable analysis, timing of vaccination in relation to transplantation, OR: 0.28 (0.15-0.54), p=0.0001; immunosuppression burden, OR: 0.22 (0.13-0.37), p<0.0001, and a diagnosis of diabetes, OR: 0.49 (0.32-0.75), p=0.001, remained independent risk factors for non-seroconversion, *Supplemental Information*, **Table S2**.

The proportion of patients who were seropositive post-V3 following receipt of the vaccine combinations ChAdOx1^2^-mRNA1273, ChAdOx1^2^-BNT162b2, BNT162b2^2^-mRNA1273 or all BNT162b2^3^, were 15/31 (48.4%), 181/245 (73.9%), 18/25 (72.0%) and 231/285 (81.1%) respectively. A significantly higher proportion of patients who had received BNT162b2 as V3 following priming with BNT162b2^2^ compared with ChAdOx1^2^ were seropositive, p=0.048. Of the 445 seropositive patients post-V3, anti-S concentrations in patients receiving ChAdOx1^2^-mRNA1273, ChAdOx1^2^-BNT162b2, BNT162b2^2^-mRNA1273 and BNT162b2^3^, were 319 (125-3213), 518 (98-2049), 412 (106-841) and 1110 (246-2969)BAU/ml respectively, with significantly higher levels in those primed with BNT162b2^2^ compared with ChAdOx1^2^, p=0.0011, *Supplemental Information*, **Figure S2**.

Following V4, 45/239 (18.8%) infection-naïve patients remained seronegative after a median period of 41 (25-64) days, **Figure 1**. De novo seroconversion post-V4 occurred in 15/60 (25.0%) patients who were seronegative post-V3. Clinical characteristics associated with lack of seroconversion post-V4 are shown in the *Supplemental Information*, **Table S3**. On multivariable analysis receiving ≥2 different classes of immunosuppression medications, OR: 0.41(0.17-0.90), p=0.033 was associated with non-seroconversion; whilst seropositivity was more likely with shorter time intervals between doses 3 and 4, OR: 0.99 (0.97-0.99), p=0.039, *Supplemental Information*, **Table S4**.

There was no difference in the proportion of patients who were seropositive post-V4 following BNT162b2^4^ compared with ChAdOx1^2^-BNT162b2^2^, at 99/115 (86.1%) versus 73/89 (82.0%) respectively, p=0.43. Serostatus post-V4 in the other vaccine combinations maybe found in the *Supplemental Information*, **Table S5**. Of the 194 seropositive patients post-V4, anti-S concentrations in patients receiving ChAdOx1^2^-BNT162b2^2^, 678 (167-284) BAU/ml, were no different compared with those patients who received BNT162b2^4^, 865 (179-3936) BAU/ml, p=0.50. Anti-S concentrations for the other vaccine combinations are shown in the *Supplemental Information*, **Table S5**.

There was a significant difference in anti-S concentration in seropositive patients post-V4 in relation to which vaccine dose led to seroconversion. The median anti-S post-V4 in infection-naïve patients who seroconverted post-V2 was 1561 (567-5211) BAU/ml, which was significantly higher than the median concentration of 379 (101-851) BAU/ml in patients who had seroconverted post-V3, p<0.0001, **Figure 2**. This was in turn significantly greater compared with those patients who only seroconverted post-V4, with a median anti-S of 19 (9.7-48) BAU/ml, p=0.0013, **Figure 2**.

**Figure 2.**
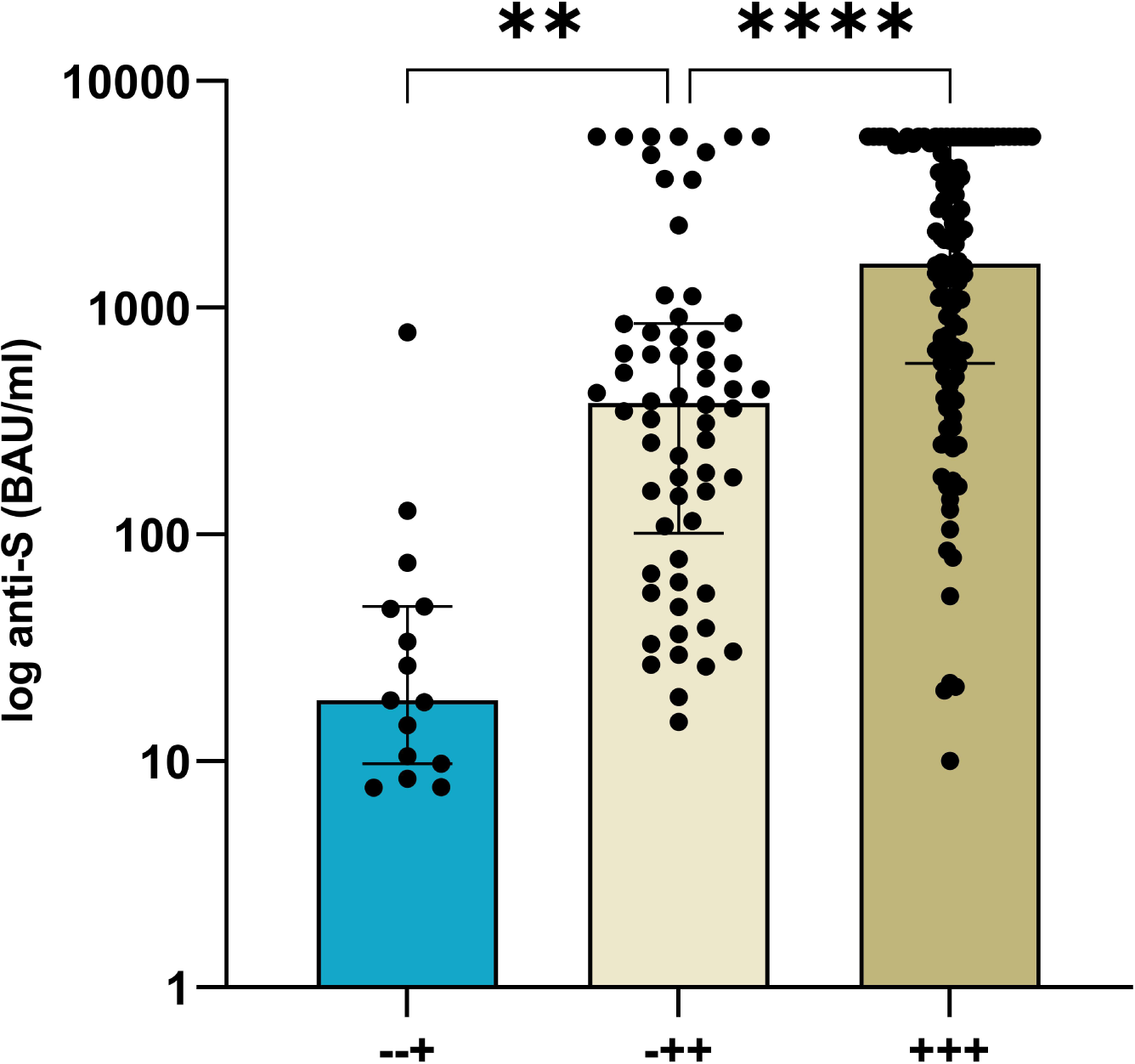
Anti-S concentrations post-V4 by dose of vaccine seroconverted. The median anti-S in infection-naïve patients post-V4 in those seroconverting post V2-, V3- or V4-were 1561 (567-5211), (101-851) and 19 (9.7-48) BAU/ml respectively.

In those patients who responded post-V2, anti-S was significantly higher post-V3, with concentrations of 148 (30-617) and 1401 (472-3213) BAU/ml respectively, p<0.0001. However, no differences were seen post-V4, 1561 (567-5211) BAU/ml, compared with post-V3, p=0.17, *Supplemental Information*, **Figure S3**.

### Comparison of anti-S concentrations by vaccination and infection status

Of 138 patients with prior infection, 6/138 (4.3%) remained seronegative post-V3 after a median time of 34 (21-48) days. Five of 6 patients had infection confirmed via RT-PCR testing, with the remaining patient having pre-vaccination positive serology. At a median time of 36 (21-59) days post-V4, 4/83 (4.8%) patients with prior infection remained seronegative, all 4 patients had infection diagnosed via RT-PCR testing.

Comparing all (seronegative and seropositive) anti-S concentrations following each vaccine, patients with a history of SARS-CoV-2 infection had significantly higher anti-S compared with infection-naïve, **Figure 3**. Post-V2 anti-S was 568 (54-2237) and 9.2 (7.1-173) BAU/ml in those with a history of SARS-CoV-2 infection compared with infection-naïve patients respectively, p<0.0001. Post-V3 concentrations in infection exposed were 3791 (1142-5680) BAU/ml compared with 295 (9.1-1611) BAU/ml in infection-naïve patients, p<0.0001; whilst post-V4 concentrations were 3993 (835-5680) and 437 (26-2211) BAU/ml respectively, p<0.0001, **Figure 3**. There was no difference between post-V2 concentrations in patients with prior exposure compared with infection-naïve individuals post-V3, p=0.06 or post-V4, p=0.99.

**Figure 3.**
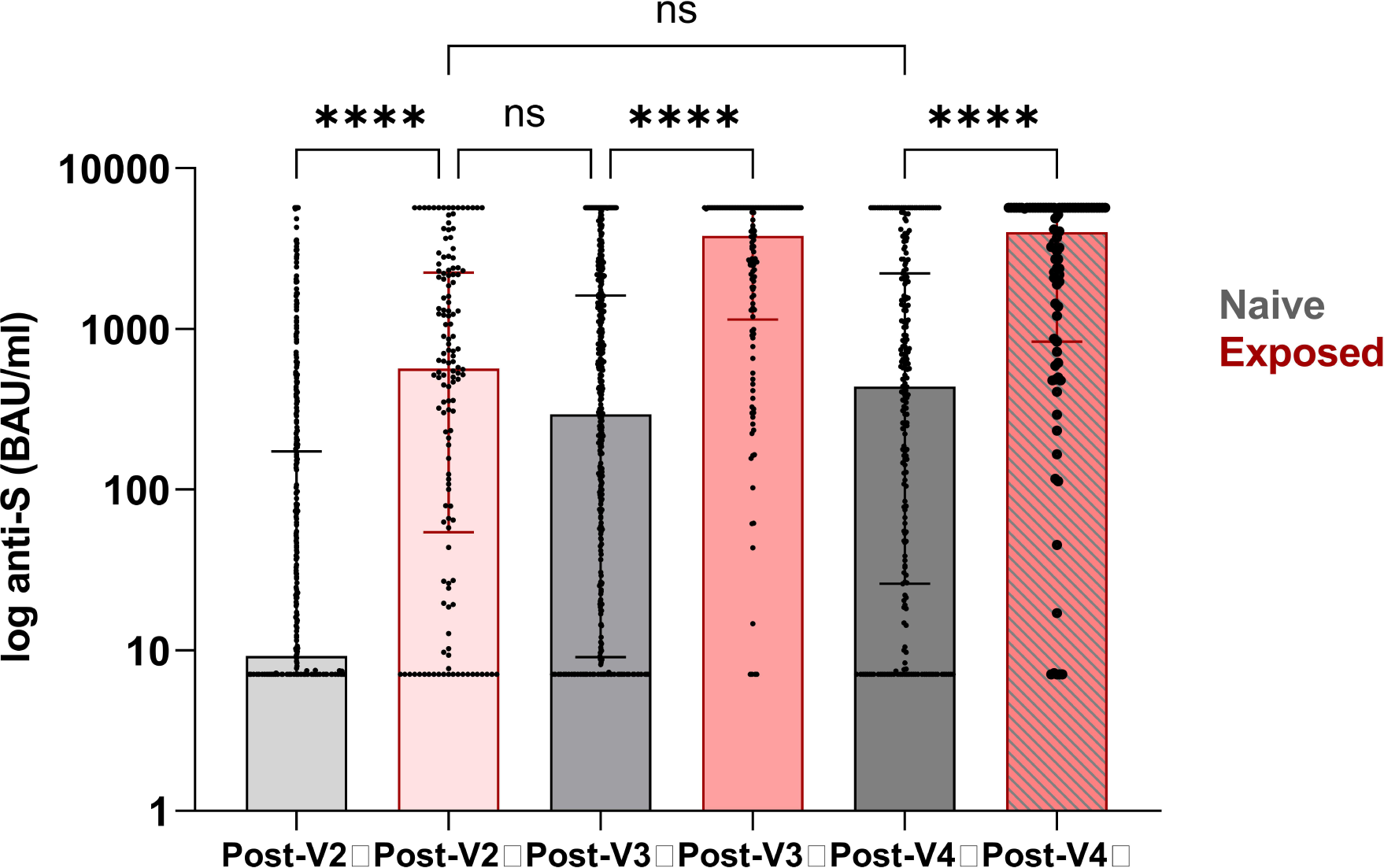
Anti-S concentrations post 2^nd^-, 3^rd^- and 4^th^ vaccinations by infection exposure. Anti-S concentrations were greater in patients with prior infection (568 (54-2237) post-V2, 3791 (1142-5680) post-V3 and 3993 (835-5680) BAU/ml post-V4) compared with infection-naïve patients (9.2 (7.1-173) post-V2, 295 (9.1-1611) post-V3 or 437 (26-2211) BAU/ml post-V4). There was no difference between post-V2 concentrations in patients with prior infection compared with infection-naïve individuals post-V3, p=0.06 or post-V4, p=0.99.

### Cellular responses post-V4

Fifty-four infection-naïve patients were assessed for T-cell responses post-V4. T-cell responses were only detectable in 11/54 (20.4%) of patients sampled at 38 (27-55) days post-V4. Clinical characteristics associated with T-cell response included younger age and being of non-Caucasian background, as shown in the *Supplemental Information*, **Table S6**. On multivariable analysis, increasing age, OR: 0.88 (0.77-0.97), p=0.026 and Caucasian ethnicity, OR: 0.03 (0.00-0.33), p=0.08, remained independent factors associated with no detectable T-cell responses.

There was no difference in the magnitude of cellular responses between those patients who were primed with ChAdOx1^2^ compared with BNT162b2^2^, with a median 9 (1-65) and 6 (2-19) SFU/10^6^ PBMC respectively, p=0.72, *Supplemental Information*, **Figure S4**. However, T-cell responses were greater in infection-naïve individuals who were seropositive, 10 (2-34) SFU/10^6^ PBMC, compared with those who were seronegative, 1 (0-8) SFU/10^6^ PBMC, **Figure 4**.

**Figure 4.**
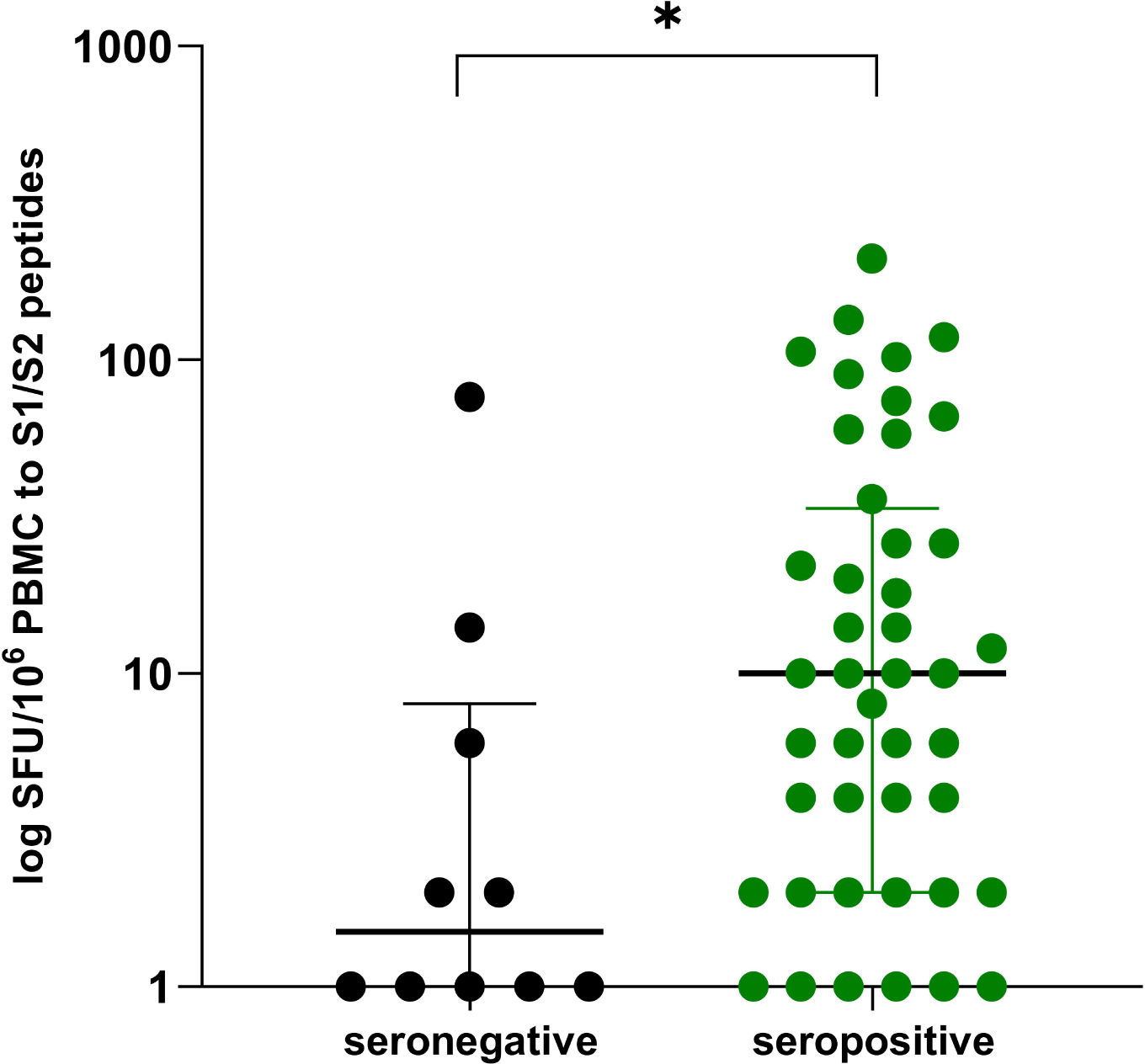
Median SFU/10^6^ PBMC by serostatus post-V4 in infection-naïve individuals. T-cell responses were greater in infection-naïve individuals who were seropositive post-V4, 10 (2-34) SFU/10^6^ PBMC, compared with those who were seronegative, 1 (0-8) SFU/10^6^ PBMC. For the purposes of the graph, 0 values were substituted by 1.

Fifteen of 83 (18.1%) infection-exposed individuals underwent T-cell assessment post-V4; 8/15 (53.3%) patients had detectable T-cell responses, which was proportionately higher than infection-naïve individuals post-V4, p=0.012. Overall T-cell responses were greater in infection exposed compared with infection-naïve individuals, with a median SFU/10^6^ PBMC of 92 (8-212) and 6 (2-26) respectively, p=0.0098, *Supplemental Information*, **Figure S5**.

## Discussion

This study shows that 24% and 19% of kidney transplant recipients do not have any detectable spike protein antibody in response to 3^rd^ and 4^th^ doses of vaccine respectively. For those patients who seroconvert after 3 or 4 doses, antibody concentrations remain lower than those patients who responded after 2 doses. Furthermore, T-cell responses are poor post-V4, which is compatible with the universal use of calcineurin inhibitors in this group of KTRs and the majority of solid organ transplant recipients across the globe. The use of such agents together with other potent immunosuppressants in transplant recipients, may also explain the weak T-cell responses to natural infection. Therefore, for solid organ transplant recipients, routine clinical testing for anti-S response will help identify those with no response, who maybe at highest risk of an adverse outcome if infected.

Three previous studies have reported on immune responses to 4 doses of mRNA-based vaccines in transplant recipients^5-7^. The total number of patients reported collectively in these studies was 122, with similar messaging that patients with no detectable anti-S response post-V3 can seroconvert post-V4 in 10-50% of cases, but they are unlikely to have significant anti-spike concentrations or possess neutralising capabilities^5,6^. However, our data are the first to include comparative V4 data in transplant recipients receiving heterologous vaccines. Given evidence suggesting that heterologous vaccination dosing may result in at least equivocal, if not enhanced, serological and cellular responses in both the general population and transplant recipients post-V3, comparing vaccination schedules is an important consideration^8,9^. After V4, we found no immune advantage of heterologous versus homologous vaccinations. Technical limitations of the study include reliance on an ELISpot assay which assesses IFN-γ as the sole read out for T-cell reactivity, rather than poly-functional cytokine responses, and lack of data on antibody neutralising capabilities^10,11^.

Consistent with this immunogenicity data, real world vaccine efficacy has been shown to be inferior in immunocompromised people, who have been at highest risk of breakthrough infections and severe disease, in the pre-Omicron era^12-14^. So, what does this mean for the strategic forward planning to protect transplant recipients? The data shown in this study suggest that a proportion of transplant recipients who have not responded to the first 4 vaccines, are unlikely to develop meaningful protection with a fifth. Whilst for other immunocompromised people, mostly those on B-cell directed therapies, robust SARS-CoV-2 T-cell responses have been demonstrated in the absence of antibodies, for solid organ transplant patients who are commonly maintained on both B-cell and T-cell inhibiting agents, this will not necessarily be the case^3,15^. With treatment options also limited in this group, related to relative contraindications and drug interactions, pre-exposure prophylaxis with passive immunity from neutralising monoclonal antibodies may currently be the best option whilst they remain effective against the current dominant variant^16^.

In summary, we have shown that repeated vaccinations will not adequately protect all transplant recipients. However, there is a spectrum of immune responses in patients in relation to vaccination and infection. It will disadvantage many immunocompromised people if they are managed as a uniform cohort irrespective of underlying disease, treatment or infection status. We recommend developing a more personalised approach to their management, starting with antibody screening which is widely available clinically to identify the vaccine non-responders who are likely to be the most immune suppressed and at risk of an adverse outcome with infection.

## Methods

### Study population

The study included 724 kidney transplant recipients, under the care of the Imperial College Renal and Transplant Centre, London. All participants were prospectively followed, and had serological testing performed following 2-(V2), and 3-(V3) vaccines, and at least once prior to their 1^st^ dose of vaccine. An additional 322 patients were investigated for immune responses following their 4^th^ dose (V4). All vaccines were received post-transplant. The study ‘The effect of COVID-19 on Renal and Immunosuppressed patients’, sponsored by Imperial College London, was approved by the Health Research Authority, Research Ethics Committee (Reference: 20/WA/0123).

### Serological testing

Serum was tested for antibodies to nucleocapsid protein (anti-NP) using the Abbott Architect SARS-CoV-2 IgG 2 step chemiluminescent immunoassay (CMIA) according to manufacturer’s instructions. This is a non-quantitative assay and samples were interpreted as positive or negative with a threshold index value of 1.4. Spike protein antibodies (anti-S IgG) were detected using the Abbott Architect SARS-CoV-2 IgG Quant II CMIA. Anti-S antibody titres are quantitative with a threshold value for positivity of 7.1 BAU/ml, to a maximum value of 5680 BAU/ml.

### Infection diagnosis

Infection was defined serologically or via confirmation with RT-PCR or lateral flow testing. The detection of anti-NP on current or historic samples, or the presence of anti-S at baseline (pre-vaccine) or historic samples, was required for the definition of prior infection by serological methods. Prior to December 2021, prior infection was determined by the presence of anti-NP or receptor binding domain (RBD) antibodies, using an in-house double binding antigen ELISA (Imperial Hybrid DABA; Imperial College London, London, UK), which detects total RBD antibodies.

### T cell ELISpot

SARS-CoV-2 specific T-cell responses were detected using the T-SPOT® Discovery SARS-CoV-2 (Oxford Immunotec) according to the manufacturer’s instructions, and as previously described^17^. In brief, peripheral blood mononuclear cells (PBMCs) were isolated from whole blood samples with the addition of T-Cell Select™ (Oxford Immunotec) where indicated. 250,000 PBMCs were plated into individual wells of a T-SPOT® Discovery SARS-CoV-2 plate. The assay measures immune responses to SARS-CoV-2 structural peptide pools; S1 protein, S2 protein, and positive PHA (phytohemagglutinin) and negative controls. Cells were incubated and interferon-γ secreting T cells were detected. Spot forming units (SFU) were detected using an automated plate reader (Autoimmun Diagnostika). Infection-naïve, unvaccinated participants were used to identify a threshold for a positive response using mean +3 standard deviation SFU/10^6^ PBMC, as previously described. This resulted in a cut-off for positivity of 40 SFU/10^6^ PBMC.

### Statistical Analysis

Statistical analysis was conducted using Prism 9.3.1 (GraphPad Software Inc., San Diego, California). Unless otherwise stated, all data are reported as median with interquartile range (IQR). Where appropriate, Mann-Whitney U and Kruskal-Wallis tests were used to assess the difference between 2 or >2 groups, with Dunn’s post-hoc test to compare individual groups. Multivariable analysis was carried out using multiple logistic regression using variables which were found to be significant on univariable analysis, p<0.05, unless otherwise stated.

## Data Availability

All data produced in the present work are contained in the manuscript

## Acknowledgments

This research is supported by the National Institute for Health Research (NIHR) Biomedical Research Centre based at Imperial College Healthcare NHS Trust and Imperial College London. The authors would like to thank the West London Kidney Patient Association, all the patients and staff at ICHNT (The Imperial COVID vaccine group and dialysis staff, and staff within the North West London Pathology laboratories). The authors are also grateful for support from The Nan Diamond Fund, Sidharth and Indira Burman, and the Auchi Charitable Foundation. MP is supported by an NIHR clinical lectureship. Work in DCT’s lab is supported by a Wellcome Trust Clinical Career Development Fellowship.

This work was performed in collaboration with the OCTAVE consortium. The OCTAVE trial, which is part of the COVID-19 Immunity National Core Study Programme, was sponsored by the University of Birmingham and funded by a grant from UK Research and Innovation (UKRI) administered by the Medical Research Council (grant reference number MC_PC_20031). It has been designated an Urgent Public Health (UPH) study by the National Institute of Health Research.

