## Supplemental Information for "Immune responses following 3^rd^ and 4^th^ doses of heterologous and homologous COVID-19 vaccines in kidney transplant recipients"

**Contents**

**Figure S1. Study flow diagram showing breakdown of sampling by post 3^rd^-dose (V3) and 4^th^-dose (V4), and infection status.**	2

### **Figure S1. Study flow diagram showing breakdown of sampling by post 3^rd^-dose (V3) and 4^th^-dose (V4), and infection status.**

Post-V4

**n=83**

Post-V4

**n=239**

Study Participants Post-V3 n=724

Infection-exposed n=138 (19.1%)

BNT162b2 Primed n=74 (53.6%)

n=44 (53.0%)

ChAdOx1 Primed n=64 (46.4%)

n=39 (47.0%)

Infection-naive n=586 (80.9%)

BNT162b2 Primed n=310 (52.9%)

n=131 (54.8%)

ChAdOx1 Primed n=276 (47.1%)

n=108 (45.2%)

### **Table S1. Clinical characteristics in 586 infection-naïve transplant recipients by serostatus following 3^rd^ primary vaccine dose**

| Characteristics | | No seroconversion  N= 141 (%) | Seroconversion  N= 445 (%) | p value |
| --- | --- | --- | --- | --- |
| Gender | Male  Female | 93 (66.0)  48 (34.0) | 291 (65.4)  154 (34.6) | 0.90 |
| Age at 1^st^ vaccine | Years (Median) | 61 (51-68) | 60 (49-67) | 0.39 |
| Ethnicity | Caucasian*  Black  Indoasian  Other | 73 (51.8)  13 (9.2)  38 (27.0)  17 (12.1) | 221 (49.7)  27 (6.1)  137 (30.8)  60 (13.5) | 0.66 |
| Cause of ESKD | Polycystic kidney disease  Glomerulonephritis*  Diabetic nephropathy  Urological  Unknown  Other | 17 (12.1)  41 (29.1)  27 (19.1)  8 (5.7)  30 (21.3)  18 (12.8) | 52 (11.7)  146 (32.8)  67 (15.1)  43 (9.7)  93 (20.9)  44 (9.9) | 0.41 |
| Number of transplants received | 1  ≥2 | 117 (83.0)  24 (17.0) | 395 (88.8)  50 (11.2) | 0.072 |
| 1^st^ vaccine <1 year post-transplant | No  Yes | 116 (82.3)  25 (17.7) | 418 (93.9)  27 (6.1) | <0.0001 |
| Type of transplant | Deceased Donor  Living Donor*  Simultaneous Pancreas-Kidney | 83 (58.9)  49 (34.8)  9 (6.4) | 240 (53.9)  193 (43.4)  12 (2.7) | 0.07 |
| Induction agent | Alemtuzumab*  IL2 receptor antagonist  None  Unknown | 76 (53.9)  32 (22.7)  5 (3.5)  28 (19.9) | 325 (73.0)  41 (9.2)  16 (3.6)  63 (14.2) | <0.0001 |
| Immunosuppression type | CNI Monotherapy*  CNI/MMF (orAza)  CNI/MMF/Prednisolone  CNI/Prednisolone  MMF (or Aza)/Prednisolone  Other | 29 (20.6)  57 (40.4)  42 (29.8)  9 (6.4)  1 (0.7)  3 (2.1) | 244 (54.8)  99 (22.2)  57 (12.8)  42 (9.4)  1 (0.2)  2 (0.4) | <0.0001 |
| Diabetes | No  Yes | 79 (56.0)  62 (44.0) | 307 (69.0)  138 (31.0) | 0.005 |
| Priming vaccine type | BNT162b2^2^  ChAdOx1^2^ | 61 (43.3)  80 (56.7) | 249 (56.0)  196 (44.0) | 0.0086 |
| Time between 1^st^ 2 vaccinations | Days (median) | 74 (63-78) | 75 (67-78) | 0.51 |
| Time between 2^nd^ -3^rd^ vaccinations | Days (median) | 167 (145-189) | 174 (156-189) | 0.033 |
| Time of serological test post-V3 | Days (median) | 24 (21-43) | 33 (21-53) | 0.007 |

*Comparator. CNI (Calcineurin inhibitor); MMF (mycophenolate); Aza (Azathioprine)

### **Table S2. Clinical characteristics associated with seroconversion following 3^rd^ primary dose vaccinations in infection naïve transplant recipients**

| **Variable** | **Reference Group** | **Univariable** | | **Multivariable** | |
| --- | --- | --- | --- | --- | --- |
|  |  | **OR (95% CI)** | **P value** | **OR (95% CI)** | **P value** |
| Induction agent | Alemtuzumab | 2.32 (1.57-3.43) | <0.0001 | 1.18 (0.74-1.87) | 0.49 |
| Immunosuppression | CNI monotherapy | 4.69 (3.03-7.46) | <0.0001 | 4.48 (2.69-7.63) | <0.0001 |
| Diabetes | Yes | 0.57 (0.39-0.85) | 0.0049 | 0.49 (0.32-0.75) | 0.001 |
| Vaccine dose 1&2 | BNT162b2 | 1.67 (1.14-2.45) | 0.0088 | 1.30 (0.85-1.98) | 0.22 |
| Vaccine within the 1^st^ year post-transplant | Yes | 0.30 (0.17-0.54) | <0.0001 | 0.28 (0.15-0.54) | 0.0001 |
| Time interval between vaccine 2&3 |  | 1.01 (1.00-1.02) | 0.007 | 1.01 (0.99-1.01) | 0.15 |
| Time to serological test |  | 1.01 (1.00-1.02) | 0.01 | 1.01 (0.99-1.02) | 0.062 |

### **Figure S2. Anti-S concentrations post-V3 in infection-naïve individuals by vaccine combination**

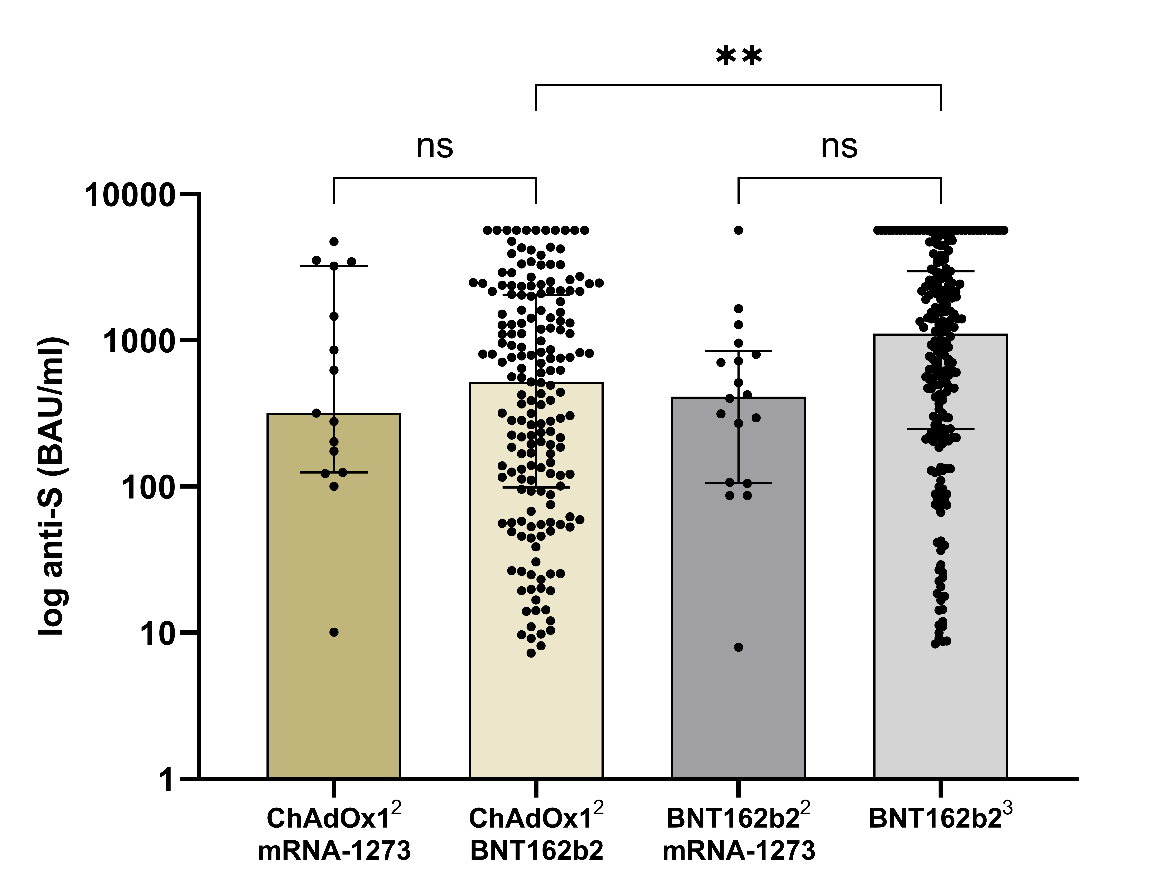

Anti-S concentrations post-V3 in infection-naïve patients receiving ChAdOx1^2^-mRNA1273, ChAdOx1^2^-BNT162b2, BNT162b2^2^-mRNA1273 and BNT162b2^3^, were 319 (125-3213), 518 (98-2049), 412 (106-841) and 1110 (246-2969) BAU/ml respectively. Significantly higher concentrations were seen with BNT162b2 as V3 following priming with BNT162b2 compared with ChAdOx1, p=0.0011

**Table S3. Clinical characteristics in 239 infection-naïve transplant recipients by serostatus following 4^th^ vaccine dose**

| Characteristics | | No seroconversion  N= 45 (%) | Seroconversion  N= 194 (%) | p value |
| --- | --- | --- | --- | --- |
| Gender | Male  Female | 27 (60.0)  18 (40.0) | 122 (62.9)  72 (37.1) | 0.72 |
| Age at 1^st^ vaccine | Years (Median) | 58 (50-66) | 61 (53-68) | 0.22 |
| Ethnicity | Caucasian  Black  Indoasian  Other | 28 (62.2)  5 (11.1)  8 (17.8)  4 (8.9) | 121 (62.4)  10 (5.2)  42 (21.6)  21 (10.8) | 0.48 |
| Cause of ESKD | Polycystic kidney disease  Glomerulonephritis  Diabetic nephropathy  Urological  Unknown  Other | 5 (11.1)  17 (37.8)  4 (8.9)  4 (8.9)  8 (17.8)  7 (15.6) | 28 (14.4)  66 (34.0)  26 (13.4)  20 (10.3)  36 (18.6)  18 (9.3) | 0.78 |
| Number of transplants received | 1  ≥2 | 32 (71.1)  13 (28.9) | 168 (86.6)  26 (13.4) | 0.012 |
| 1^st^ vaccine <1 year post-transplant | No  Yes | 39 (86.7)  6 (13.3) | 183 (94.3)  11 (5.7) | 0.07 |
| Type of transplant | Deceased Donor  Living Donor  Simultaneous Pancreas-Kidney | 25 (55.6)  17 (37.8)  3 (6.7) | 92 (47.4)  93 (47.9)  9 (4.6) | 0.45 |
| Induction agent | Alemtuzumab*  IL2 receptor antagonist  None  Unknown | 25 (55.6)  12 (26.7)  0  8 (17.8) | 123 (63.4)  25 (12.9)  13 (6.7)  33 (17.0) | 0.049 |
| Immunosuppression type | CNI Monotherapy*  CNI/MMF (orAza)  CNI/MMF/Prednisolone  CNI/Prednisolone  MMF (or Aza)/Prednisolone  Other | 9 (20.0)  11 (24.4)  21 (46.7)  3 (6.7)  -  1 (2.2) | 88 (45.4)  64 (33.0)  20 (10.3)  19 (9.8)  1 (0.5)  2 (1.0) | <0.0001 |
| Diabetes | No  Yes | 30 (66.7)  15 (33.3) | 136 (70.1)  58 (29.9) | 0.65 |
| Vaccine type | BNT162b2  ChAdOx1 | 20 (44.4)  25 (55.6) | 111 (57.2)  83 (42.8) | 0.12 |
| Vaccine combination | BNT162b2/ mRNA-1273  BNT162b2/ mRNA-1273/ BNT162b2  BNT162b2  ChAdOx1/mRNA-1273  ChAdOx1/mRNA-1273/ BNT162b2  ChAdOx1/ BNT162b2 | 1 (2.2)  3 (6.7)  16 (35.6)  5 (11.1)  4 (8.9)  16 (35.6) | -  12 (6.4)  99 (52.9)  2 (1.1)  8 (4.3)  73 (37.6) | 0.001 |
| Time between 1^st^ and 2^nd^ vaccinations | Days (median) | 71 (63-77) | 75 (65-78) | 0.21 |
| Time between 2^nd^ and 3^rd^ vaccinations | Days (median) | 160 (142-189) | 167 (153-184) | 0.41 |
| Time between 3^rd^ and 4^th^ vaccinations | Days (median) | 116 (95-130) | 98 (92-112) | 0.005 |
| Time of serological test post-V4 | Days (median) | 38 (28-53) | 42 (23-66) | 0.66 |

### **Table S4. Clinical characteristics associated with seropositivity following 4^th^ primary dose vaccinations in infection-naïve transplant recipients**

| **Variable** | **Reference Group** | **Univariable** | | **Multivariable** | |
| --- | --- | --- | --- | --- | --- |
|  |  | **OR (95% CI)** | **P value** | **OR (95% CI)** | **P value** |
| Number of transplants | Two | 0.38 (0.18-0.84) | 0.014 | 0.47 (0.21-1.07) | 0.06 |
| Immunosuppression | CNI monotherapy | 3.32 (1.58-7.68) | 0.0027 | 2.44 (1.11-5.80) | 0.033 |
| Priming (dose 1&2) vaccine type | BNT162b2 | 1.67 (0.87-3.24) | 0.12 | 1.31 (0.65-2.64) | 0.45 |
| Vaccine within the 1^st^ year post-transplant | Yes | 0.39 (1.4-1.19) | 0.08 | 0.57 (0.19 -1.87) | 0.33 |
| Time interval between vaccine doses 3&4 | Days | 0.98 (0.97-0.99) | 0.005 | 0.99 (0.97-0.99) | 0.039 |

### **Table S5. Proportion of infection-naïve transplant patients with seropositive status and anti-S concentrations post-V4 by vaccine type**

| **Vaccine combination** | **Total number of patients**  **n=239 (%)** | **Seronegative** | **Seropositive** | **Median anti-S concentrations in**  **seropositive patients (BAU/ml)** |
| --- | --- | --- | --- | --- |
| ChAdOx1^2^-mRNA1273^2^ | 7 (2.9) | 5 (71.4) | 2 (28.6) | 971 (857-1086) |
| ChAdOx1^2^-mRNA1273-BNT162b2 | 12 (5.0) | 4 (33.3) | 8 (66.7) | 637 (38-4454) |
| ChAdOx1^2^-BNT162b2^2^ | 89 (37.2) | 16 (18.0) | 73 (82.0) | 678 (167-284) |
| BNT162b2^2^- mRNA1273^2^ | 1 (0.4) | 1 (100) | - | - |
| BNT162b2^2^- mRNA1273- BNT162b2 | 15 (6.3) | 3 (20.0) | 12 (80.0) | 478 (182-4888) |
| BNT162b2^4^ | 115 (48.1) | 16 (13.9) | 99 (86.1) | 865 (179-3936) |
| **Total** | **239** | **45** | **194** | 741 (178-3495) |

### **Figure S3. Anti-S concentrations post-V3 and post-V4 in patients who seroconverted post-V2**

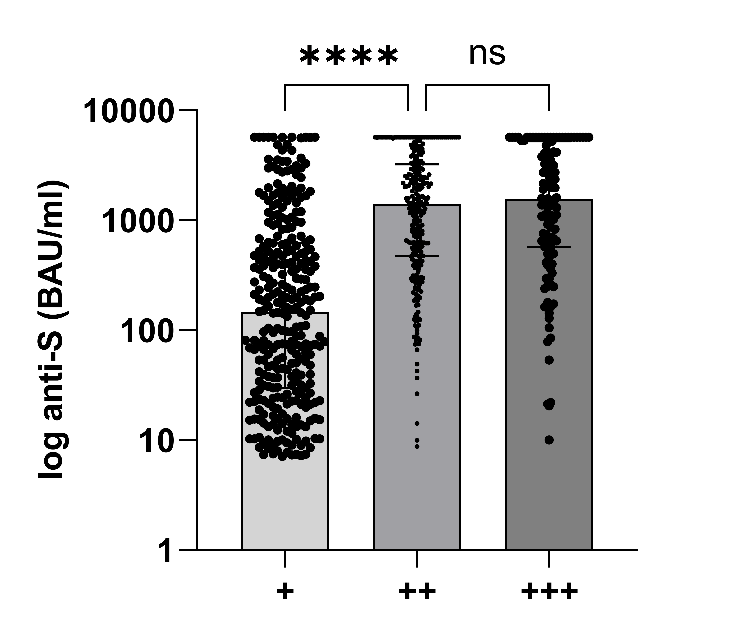

Anti-S concentrations post-V2, V3 and V4 in patients who seroconverted post-V2 were 148 (30-617), 1401 (472-3213) and 1561 (567-5211) BAU/ml respectively. Post-V2 compared with post-V3, p<0.0001, and post-V3 compared with post-V4, p=0.17.

### **Table S6. Clinical characteristics in 54 infection-naïve transplant recipients by T-cell response following 4th vaccine dose**

| Characteristics | | No T-cell response  N= 43 (%) | T-cell response  N= 11 (%) | p value |
| --- | --- | --- | --- | --- |
| Gender | Male  Female | 30 (69.8)  13 (30.2) | 7 (63.6)  4 (36.4) | 0.70 |
| Age at 1^st^ vaccine | Years (Median) | 62 (55-78) | 50 (42-59) | 0.006 |
| Ethnicity | Caucasian*  Black  Indoasian  Other | 23 (53.5)  2 (4.7)  11 (25.6)  7 (16.3) | 2 (18.2)  2 (18.2)  3 (27.3)  4 (36.4) | 0.038 |
| Cause of ESKD | Polycystic kidney disease  Glomerulonephritis*  Diabetic nephropathy  Urological  Unknown  Other | 6 (14.0)  13 (30.2)  5 (11.6)  6 (14.0)  8 (18.6)  5 (11.6) | -  6 (54.5)  1 (9.1)  -  3 (27.3)  1 (9.1) | 0.42 |
| Number of transplants received | 1  ≥2 | 40 (93.0)  3 (7.0) | 8 (72.7)  3 (27.3) | 0.058 |
| 1^st^ vaccine <1 year post-transplant | No  Yes | 40 (93.0)  3 (7.0) | 9 (81.8)  2 (18.2) | 0.26 |
| Type of transplant | Deceased Donor  Living Donor*  Simultaneous Pancreas-Kidney | 16 (37.2)  24 (55.8)  3 (7.0) | 5 (45.5)  6 (54.5)  - | 0.94 |
| Induction agent | Alemtuzumab*  IL2 receptor antagonist  None  Unknown | 30 (69.8)  7 (16.3)  0  6 (14.0) | 5 (45.5)  1 (9.1)  1 (9.1)  4 (36.4) | 0.14 |
| Immunosuppression type | CNI Monotherapy*  CNI/MMF (orAza)  CNI/MMF/Prednisolone  CNI/Prednisolone  MMF (or Aza)/Prednisolone  Other | 16 (37.2)  13 (30.2)  10 (23.3)  4 (9.3)  -  - | 3 (27.3)  3 (27.3)  1 (9.1)  4 (36.4) | 0.54 |
| Diabetes | No  Yes | 27 (62.8)  16 (37.2) | 9 (81.8)  2 (18.2) | 0.24 |
| Vaccine type | BNT162b2  ChAdOx1 | 26 (60.5)  17 (39.5) | 4 (36.4)  7 (63.6) | 0.16 |
| Vaccine combination | BNT162b2/ mRNA-1273  BNT162b2/ mRNA-1273/ BNT162b2  BNT162b2  ChAdOx1/mRNA-1273  ChAdOx1/mRNA-1273/ BNT162b2  ChAdOx1/ BNT162b2 | 1 (2.3)  5 (11.6)  20 (46.5)  2 (4.7)  2 (4.7)  13 (30.2) | -  1 (9.1)  3 (27.3)  -  -  7 (63.6) | 0.45 |
| Seroconversion | No  Yes | 9 (20.9)  34 (79.1) | 1 (9.1)  10 (90.9) | 0.37 |
| Time between 1^st^ and 2^nd^ vaccinations | Days (median) | 75 (66-78) | 68 (63-74) | 0.20 |
| Time between 2^nd^ and 3^rd^ vaccinations | Days (median) | 156 (143-178) | 155 (151-179) | 0.76 |
| Time between 3^rd^ and 4^th^ vaccinations | Days (median) | 103 (92-126) | 102 (95-114) | 0.81 |
| Time of testing post-V4 | Days (median) | 56 (27-81) | 30 (22-37) | 0.046 |

### **Table S7. Multivariate analysis of clinical characteristics associated with T-cell response following 4^th^ primary dose vaccinations in infection naïve transplant recipients**

| **Variable** | **Reference Group** | **Univariable** | | **Multivariable** | |
| --- | --- | --- | --- | --- | --- |
|  |  | **OR (95% CI)** | **P value** | **OR (95% CI)** | **P value** |
| Age |  | 0.92 (0.85-0.97) | 0.009 | 0.88 (0.77-0.97) | 0.026 |
| Induction agent | Alemtuzumab | 0.36 (0.09-1.40) | 0.14 | 0.59 (0.09-3.85) | 0.57 |
| Ethnicity | White | 0.19 (0.03-0.86) | 0.05 | 0.03 (0.00-0.33) | 0.018 |
| Graft number | 1^st^ | 0.2 (0.03-1.25) | 0.075 | 0.04 (0.00-0.46) | 0.026 |
| Time of testing post-V4 |  | 0.96 (0.91-0.99) | 0.05 | 0.96 (0.90-1.01) | 0.17 |

### **Figure S4. T-cell responses in 54 infection-naïve patients post-V4 by priming vaccine type***

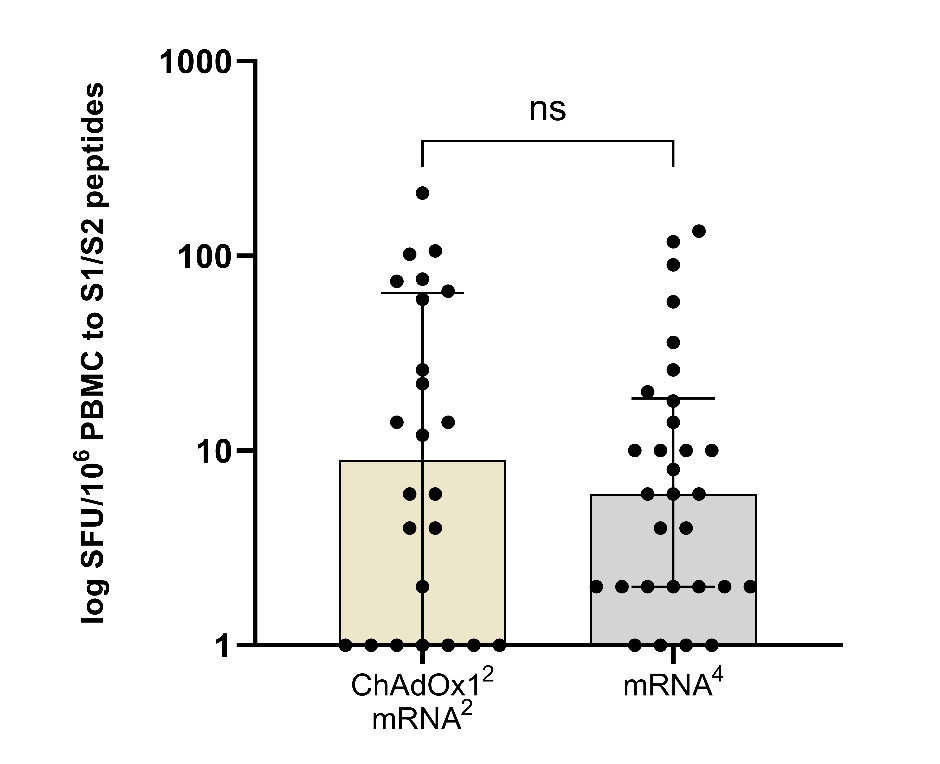

There was no difference in the magnitude of cellular responses between those patients who were primed with ChAdOx1^2^ compared with BNT162b2^2^, with a median 9 (1-65) and 6 (2-19) SFU/10^6^ PBMC respectively, p=0.72. *For purposes of graphical representation, values of 0 were replaced by 1.

### **Figure S5. T-cell responses in infection-naïve versus infection exposed patients post-V4***

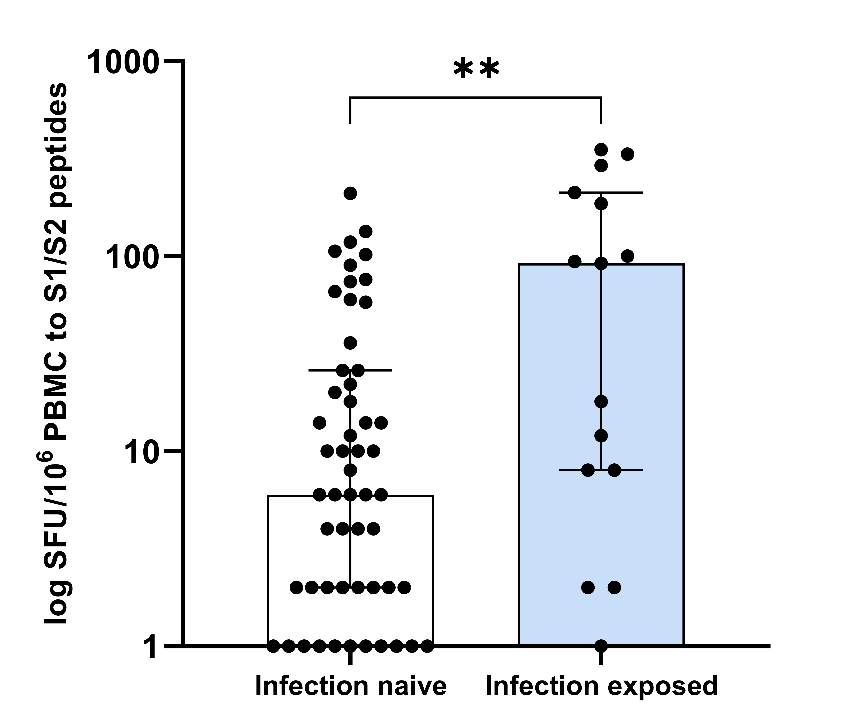

T-cell responses were greater in infection exposed compared with infection-naïve individuals, with a median SFU/10^6^ PBMC of 92 (8-212) and 6 (2-26) respectively, p=0.0098. *For purposes of graphical representation, values of 0 were replaced by 1.
